# A controlled clinical effectiveness trial of multimodal cognitive rehabilitation on episodic memory functioning in older adults with traumatic brain injury (TBI)

**DOI:** 10.1101/2020.10.26.20216523

**Authors:** Eduardo Cisneros, Elaine de Guise, Sylvie Belleville, Michelle McKerral

## Abstract

**Objectives:** This study aimed to evaluate the impact of a multimodal cognitive intervention, the Cognitive Enrichment Program (CEP), on episodic memory in traumatic brain injured (TBI) older adults, as compared to an active control group that received usual care in the form of holistic rehabilitation.

**Methods:** The CEP’s Memory module consisted in memory strategies to promote encoding. Effectiveness was evaluated by psychometric tests (Face-name association, Word list recall, Text memory), while generalization was measured through self-reported questionnaires about daily memory functioning (Self-Evaluation Memory Questionnaire) and psychological well-being (Psychological General Well-Being Index). Measures were obtained before and after intervention, and six months later.

**Results:** Both groups showed improvement on most measures, but the experimental group showed greater statistically significant improvement. ANCOVA mixed model repeated measures analysis showed a strong group-by-time interaction for the *Face-name association* test, with a large effect size. A significant group-by-time interaction was obtained on three generalization self-report measures, including increased memorization of the content of *Conversations*, reduced *Slips of attention*, and increased memory of *Political & social Events*, with moderate to large effect sizes. Clinically significant improvements were found for *Psychological well-being* in the experimental group, where 50% of participants improved to the well-being category and remained stable six months later (9/17; 53%). Also, the number of experimental participants showing severe distress before CEP training (9) was reduced after intervention (5) and remained relatively stable at 6 months.

**Conclusions:** The CEP is a promising cognitive rehabilitation program that showed high satisfaction in participants and that can not only improve episodic memory in terms of psychometric scores, but also in daily life situations, as well as enhance psychological well-being in older individuals having sustained a TBI.

## Introduction

Traumatic brain injury (TBI) is defined as an alteration in brain function or other evidence of brain pathology caused by external forces [1]. TBI in older adults has become a significant health problem because its incidence rises following an exponential curve as people get older, and because in a worldwide context of increasing population aging, the occurrence of TBI in older people is significantly rising [2]. However, cognitive rehabilitation programs adapted for older TBI individuals are seemingly inexistent.

Post-TBI alterations in brain functions result in a broad spectrum of cognitive, affective, behavioral and psychosocial disorders which, combined with existing cognitive characteristics of normal aging, will increase cognitive vulnerability [3]. Indeed, memory problems are the more common and pervasive complaints among individuals with TBI and are as frequent in seniors [4,5]. TBI may thus worsen memory dysfunctions that could be already present in normal aging, such as short-term episodic memory, working memory, and attention and memory processes related to cognitive control or regulation. Memory deficits can have significant negative impacts on daily activities and psychosocial independence, particularly in older people who already show physical, cognitive and psychosocial vulnerabilities [6]. On the other hand, semantic and implicit memory are generally spared in both normal aging as well as after TBI [7].

A review of RCTs of cognitive interventions in healthy older adults and in mild cognitive impairment (MCI) showed that cognitive rehabilitation can be effective in improving various aspects of cognitive functioning as reflected in objectively measured attention and memory performance, executive functioning, processing speed, fluid intelligence, as well as in self-reported cognitive performance. The heterogeneity and nature of the reviewed studies make it, however, difficult to establish if there was generalization of these gains to everyday life [8].

In terms of memory training techniques, visual imagery, strategic memory methods, method of loci, semantic associations, spaced retrieval, categorization and other specific memory techniques have demonstrated effectiveness for improving cognitive performance in healthy older adults [9,10]. Stability of such effects over time has been also demonstrated [11]. Some of those methods have been successfully applied to neuropathological populations as MCI. Belleville and collaborators [12] developed a comprehensive memory program (MEMO) for adults with MCI, which focuses on attention and encoding self-initiated strategies based on visual imagery for face-name associations and word lists, and on semantic analysis and synthesis of texts. They showed that MEMO training significantly improved delayed recall of word lists, and face-name associations, but not of memory of texts, as well as self-reported daily memory functioning and psychological general well-being.

In cognitive rehabilitation programs developed for younger adults with TBI, emphasis has been put on training of self-regulated aspects of memory, which is probably disturbed by executive dysfunctions. For example, Shefft et al. [13] tested a self-generated memory encoding strategy requiring active creation of new visual images to improve verbal learning in young persons with TBI; they demonstrated improvement on recognition tasks and cued recall, but not on free recall. Shum et al. [14] conducted a RCT to assess efficacy of four intervention modalities: self-awareness training plus compensatory prospective memory training, self-awareness training plus active control, active control plus compensatory prospective memory training, and an active control only. The two groups that received compensatory prospective memory training showed significantly higher prospective memory scores and self-reported daily strategy use.

To our knowledge, there is to date no published comprehensive cognitive rehabilitation program designed for older TBI individuals and addressing, among others, episodic memory functioning. This paper reports on the memory component of a newly designed cognitive rehabilitation program, the Cognitive Enrichment Program (CEP), which was tailored for individuals who sustain a TBI in later adulthood. The CEP is a 12-week multimodal intervention structured into three modules designed to simultaneously address cognitive problems resulting from TBI, as well as age-related cognitive issues in the following domains: self-awareness, attention and episodic memory, and executive functions. The memory module integrates training on self-initiated memory strategies based on visual imagery and semantic associations, as well as their use in real-world situations.

We evaluated the effectiveness of the CEP for enhancing episodic memory in older adults with TBI, as assessed with three neuropsychological memory measures as primary outcomes, and a questionnaire of daily memory functioning as a generalization measure of real-world memory performance. We hypothesized that self-initiated encoding training included in the CEP would result in an improvement in both psychometric and self-reported memory scores in a trained group of older individuals with TBI, whereas this would not be the case for a comparable TBI group who did not receive the CEP intervention. Secondary objectives were to evaluate more global generalization effects of CEP memory training on the psychological well-being of participants and their satisfaction as to the positive impacts of the intervention on their daily life, as well as to assess stability of change six months after the end of training.

## Material and Methods

### Experimental design

Results presented in this paper are part of a larger semi-randomized, controlled, before-after study with follow-up at 6 months and blinded outcome measurement clinical trial (study registered at ClinicalTrials.gov, Identifier: NCT04590911). This study is reported according to the TREND statement [15].

### Participants

42 French-speaking participants having sustained a TBI (based on the WHO criteria [16]) and meeting the inclusion criteria (see Supplementary material for inclusion/exclusion criteria) were recruited by clinical coordinators from the McGill University Hospital Centre (MUHC), as well as from two outpatient interdisciplinary rehabilitation centres, the Lucie-Bruneau Rehabilitation Centre and Le Bouclier Rehabilitation Centre. Ethical Review Boards of the MUHC as well as from the Centre for Interdisciplinary Research in Rehabilitation to which are affiliated the referring rehabilitation centres provided ethical approval for the study. All participants gave their written informed consent prior to their participation in the study. The study was conducted in compliance with the Helsinki Declaration. A symbolic financial compensation was offered to all participants as a contribution towards their transportation expenses. The CONSORT diagram illustrating participant allocation and flow throughout the trial is presented in Figure 1. Additional details on participant recruitment and allocation are presented as Supplementary material.

**Figure 1.**
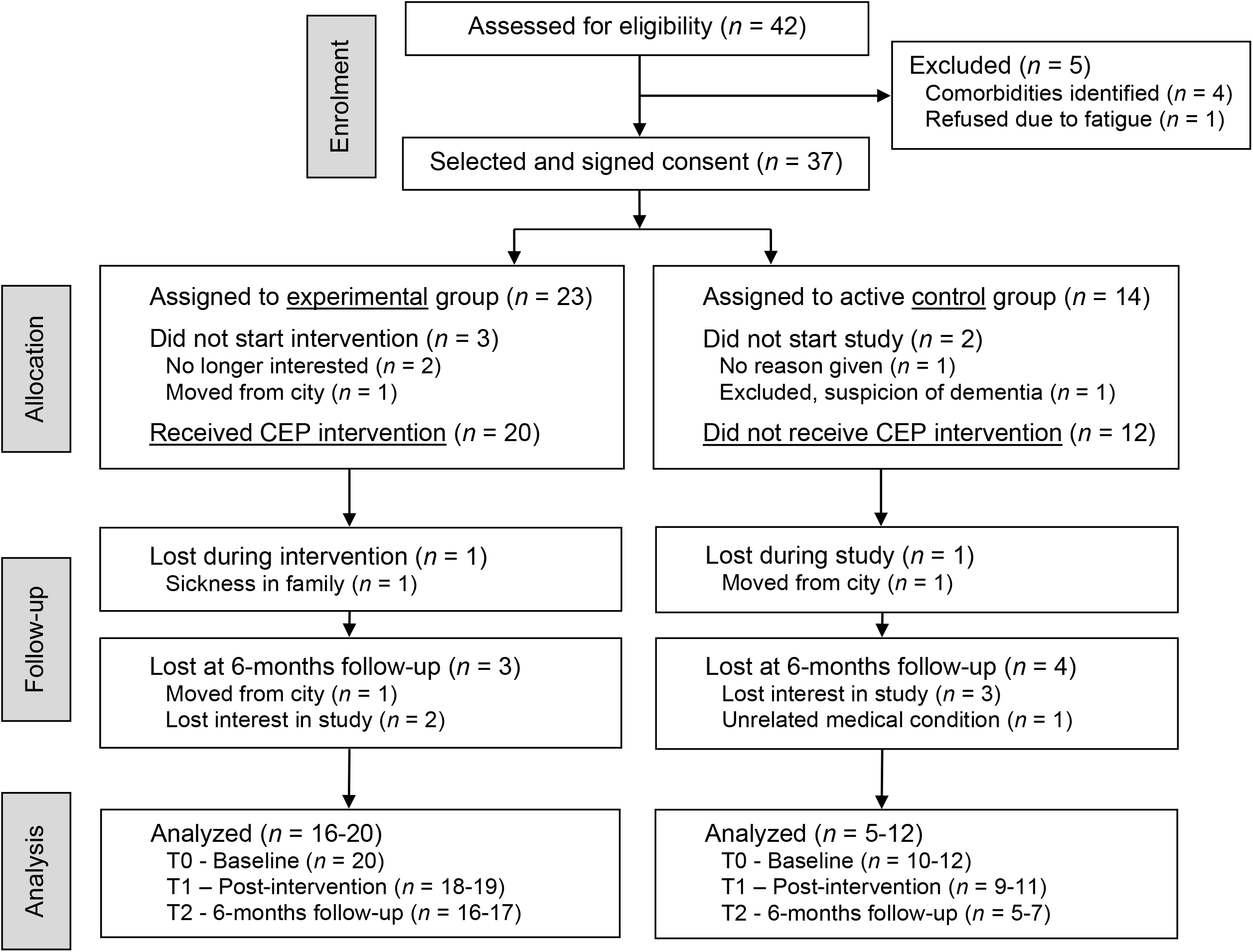
CONSORT Flow Diagram, modified for partially randomized trial design.

### Procedure

The CEP consists of three intervention modules, Introduction and self-awareness, Attention and memory, and Executive functions (strategies detailed in Cisneros et al. [17]; this issue), which include a total of 24 sessions of 90 minutes, delivered twice weekly during in a 12-week time frame. CEP main content and structure, as well as detailed intervention delivery, are presented as Supplementary material. The module targeting memory consists of the MEMO program strategies that have been described elsewhere [12]. MEMO teaches participants to use self-initiated strategies in situations requiring sustained attention and episodic memory. The main goal is to enhance and enrich encoding, and facilitate ulterior recall, by generating new visual images (method of loci, face-name associations), or by analyzing and synthesizing the semantic content of texts (FRST method). In addition to MEMO’s proposed exercises, homework included reading magazines or newspapers articles to practice the FRST method, meeting at least one new person during the week to practice the Face-name method and making one grocery list and one to-do list during the weekend. The CEP was conducted by an experienced clinical neuropsychologist (E.C., experimenter), who had previously received MEMO training, with small groups of 5 participants. Assessments of intervention effects were performed by trained evaluators, who were blinded to group assignment of participants, in two separate sessions lasting about 90 minutes each. There were three time-points: baseline (pre-intervention – T0); 14-weeks (post-intervention – T1); 6-months post-intervention (follow-up – T2). Additional information on assessments can be found in Supplementary material.

### Primary outcome measures

Three tasks were chosen as primary outcomes for episodic memory. The same tests as Belleville et al. [12] were used to evaluate the effects of the MEMO program were administered: ***Face-name association, Word list recall***, and ***Text memory***. These tests were selected from the *Côte-des-Neiges Computerized Memory Battery* [18]. See Supplementary material for more details on the measures used.

### Generalization measures

To assess the participant’s perception of changes in memory abilities in daily life after the CEP intervention, we used a self-administered memory questionnaire, the ***Self-Evaluation Memory Questionnaire (SEMQ)*** [19]. The SEMQ measures memory performance for 10 dimensions for which are derived mean scores: ***Conversations, Books & movies, Slips of attention, People, Use of objects, Political & social events, Places, Actions to perform, Personal events, General*** (this latter dimension assessing the impact of physical, emotional or environmental variables on memory). We also used the French version of the ***Psychological General Well-Being Index*** (***PGWBI)*** [20], and a ***Satisfaction questionnaire*** asking about the perceived impacts of strategies learned in each CEP module and, overall, on daily life activities.

### Control measures

The following measures served as a control of spontaneous recovery. We did not expect significant changes on these measures in any of the groups. ***Forward and Backward Digit spans*** and ***Coding*** subtests (scaled scores) from the WAIS-III were used. The *Montreal Cognitive Assessment (MoCA)* was used as a control measure for post-TBI cognitive decline associated with aging [21]. WAIS-III *Vocabulary* was used as a control measure for dementia suspicion or low intellectual functioning. As a part of general health variables describing the sample, we documented *Vascular risk factors* of each participant (e.g., arterial hypertension, cardiovascular disease) and family history of these conditions using a health questionnaire.

### Statistical analysis

Demographic, clinical characteristics, primary outcomes, generalization and control measures at baseline (T0) were described for each group and compared using Student’s t-test and chi-squared test. Since there was internal variability in both groups in time since injury and the latter could be related to enhanced time-related brain atrophic changes [22,23], we used a mixed model ANCOVA for repeated measures with a repeated factor time (T0 – pre-intervention baseline, T1 – post-intervention, T2 – 6-months follow-up), a factor group (experimental, control) and *Time since TBI* as a covariate. In this model, the effect of the intervention corresponds to a significant group-by-time interaction. Appropriate post-hoc tests were conducted. Effect sizes were interpreted according to Cohen, as cited by Pallant [24]. Appropriate post hoc comparisons were performed where indicated in this model. Analyses were performed assuming missing data are at random (MAR) with SAS version 9.4 and a significance level of *p* < .05.

## Results

Demographic and clinical characteristics of the two groups are summarized in Table 1. There were no significant differences between experimental and control groups for all demographic and clinical variables, except for marital status. The vast majority of participants in the experimental group were married or in a common-law partnership (90%), while 41.7% of control participants had a partner at time of study. Twenty-one (65%) participants integrated the research with a diagnosis of uncomplicated (8) or complicated (13) mild TBI, the latter being defined as a mild TBI with positive non-contrast CT scan. Six participants had a moderate TBI, and five presented a severe TBI.

**Table 1.** Means (*SD*) or *n* (%) for demographic and clinical variables.

|  | Experimental group<br>(*n* = 20) | Control group<br>(*n* = 12) | df | Pearson<br>Chi-square | *t* | *p* |
| --- | --- | --- | --- | --- | --- | --- |
| **Age** (years) | 64.90 (7.18) | 63.75 (5.63) | 30 |  | 0.473 | 0.639 |
| **Sex** (M/F) | 14/6 (70/30) | 5/7 (41.7/58.3) | 1 | 2.50 |  | 0.150 |
| **Education** (years) | 12.30 (2.66) | 11.83 (2.98) | 30 |  | 0.460 | 0.649 |
| **Marital Status** |  |  | 4 | 13.04 |  | 0.004 * |
| Married | 13 (65.0) | 5 (41.7) |  |  |  |  |
| Civil union | 5 (25.0) | 0 (0.0) |  |  |  |  |
| Separated | 1 (5.0) | 0 (0.0) |  |  |  |  |
| Widower | 0 (0.0) | 2 (16.6) |  |  |  |  |
| Single | 1 (5.0) | 5 (41.7) |  |  |  |  |
| **TBI severity** |  |  | 3 | 5.81 |  | 0.144 |
| Mild | 6 (30.0) | 2 (16.7) |  |  |  |  |
| Complicated mild | 8 (40.0) | 5 (41.7) |  |  |  |  |
| Moderate | 3 (15.0) | 3 (25.0) |  |  |  |  |
| Severe | 3 (15.0) | 2 (16.7) |  |  |  |  |
| **Time since TBI** (days) | 595.75 (926,67) | 859.33 (772,04) | 30 |  | -0.827 | 0.415 |
| **MoCA** (/30) | 25.90 (1.92) | 24.83 (3.27) | 30 |  | 1.169 | 0.252 |
| **Vocabulary** (scaled score) | 10.20 (1.64) | 9.25 (2.90) | 30 |  | 1.190 | 0.243 |
| **Number of vascular risk factors** | 1.93 (1.76) | 2.88 (2.02) | 30 |  | -1.396 | 0.173 |

\* Statistically significant result; MoCa: Montreal Cognitive Assessment.

### Primary outcome measures

Before intervention (T0), experimental and control groups did not present significant differences on all psychometric variables, except for macrostructure immediate recall on text memory (*t*(30) = 2.30, *p* = 0.028) where experimental participants showed higher scores than controls. Table 2 summarizes means and SD at the three assessment times, and ANCOVA group-by-time interaction results after controlling for the effects of the covariate *Time since TBI*. The analysis showed a strong significant group-by-time interaction for ***Face-name association***, with a large effect size. Supplementary bilateral t-test analysis indicated that scores at T1 were significantly larger for the experimental group than for controls (*t*(28) = 2.95, *p* = 0.006; 95% CI [1.05, 5.81]). The pre-post-intervention difference for the experimental group was also significant (*t*(28) = 2.389, *p* = 0.024; 95% CI [0.35, 4.62]). These significant differences were, however, not maintained at the 6-months follow-up, even though scores remained higher in the experimental group. Statistics for non-significant results not presented in Table 2 for and completed/attrition participants are presented as Supplementary material.

**Table 2.**
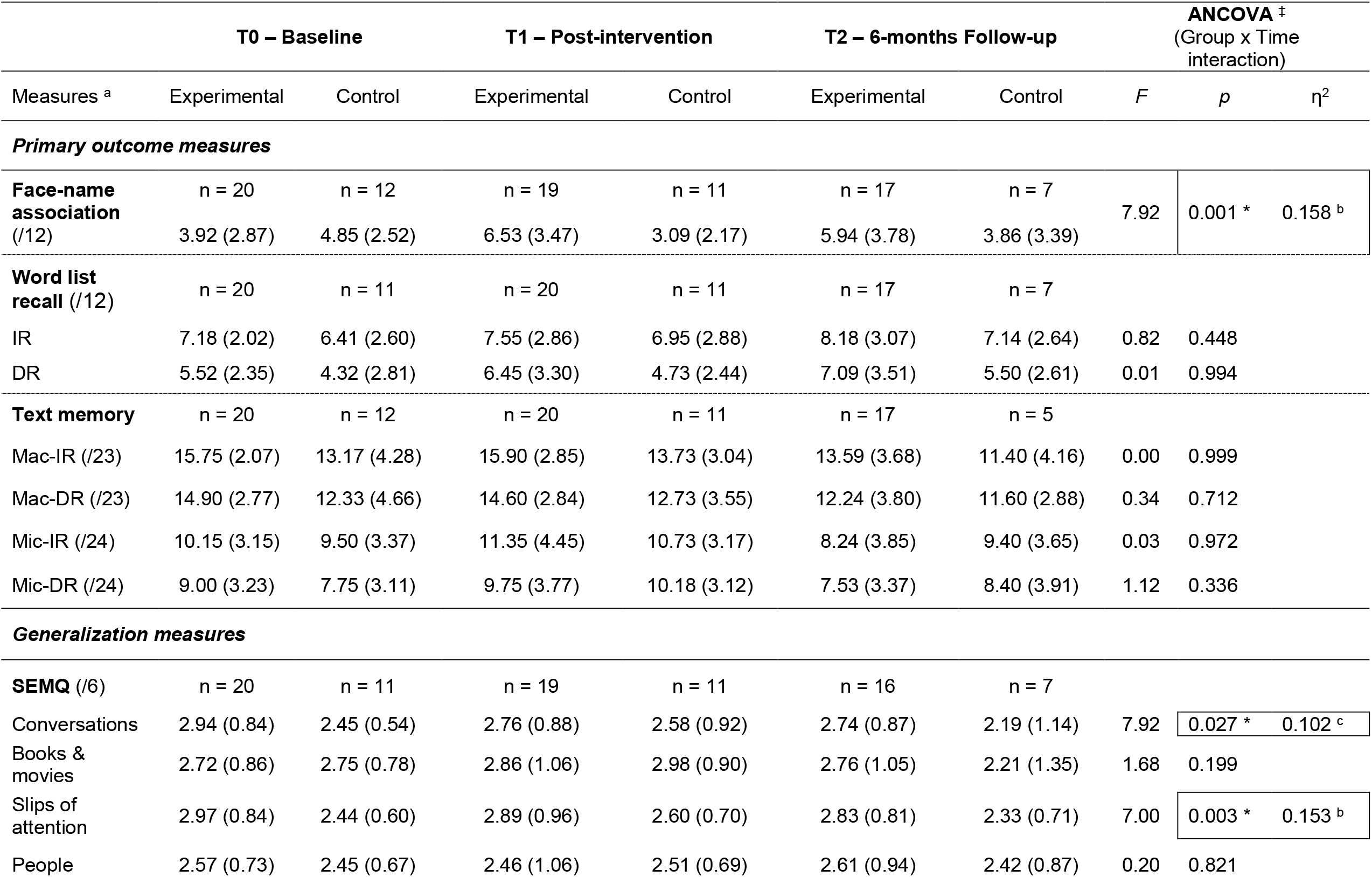
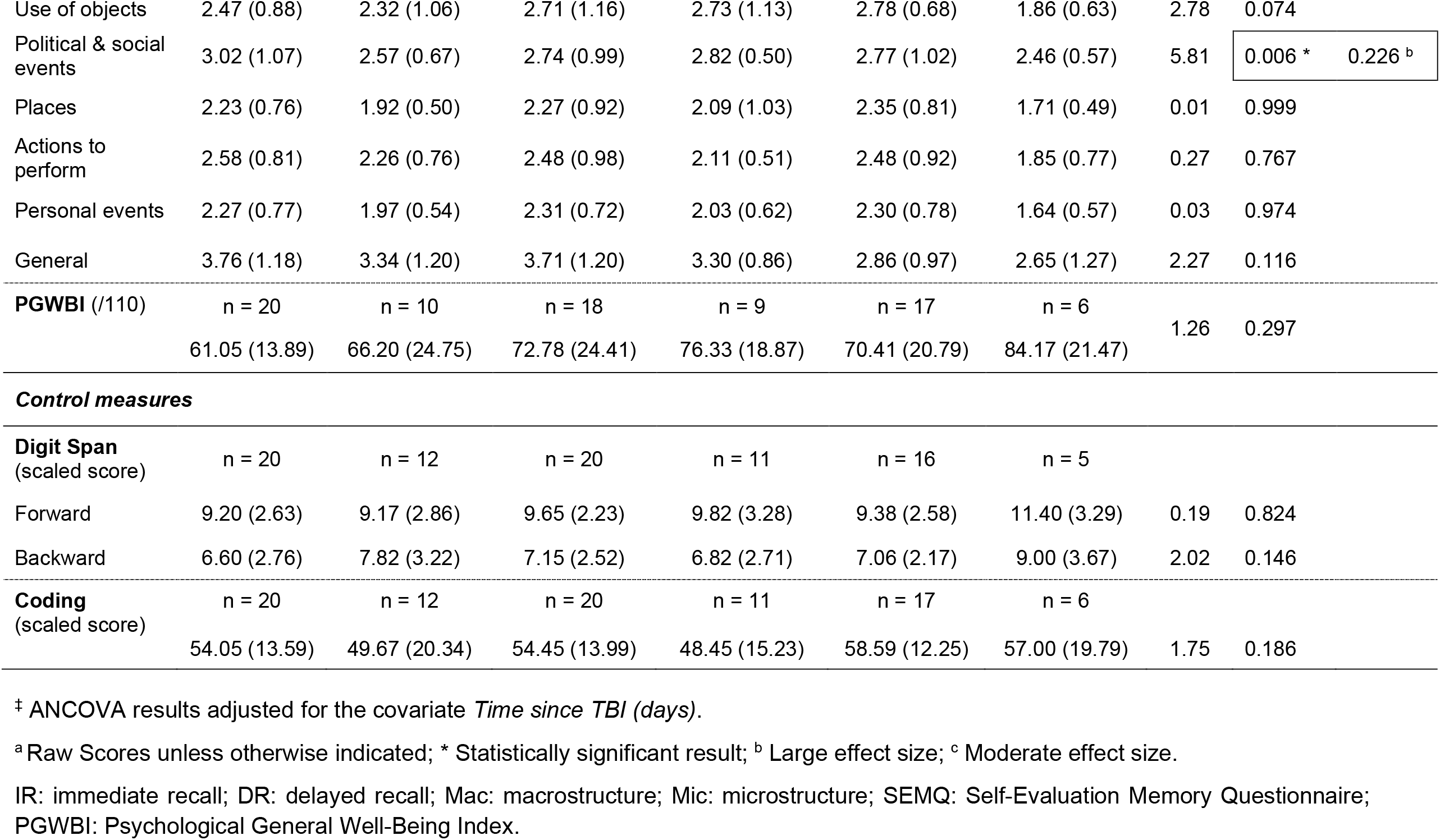
Means *(SD)* for the different measures at the three assessment times, and ANCOVA results.

There was a significant main effect of time on ***Word list - immediate recall*** (*F*(2, 44) = 3.60, *p* = 0.036), as well as for ***Word list - delayed recall*** (*F*(2, 42) = 9.23, *p* = 0.001), but there were no significant effects for group or group x time interaction. There was no significant main effect of time, group, nor a group-by-time interaction for ***Text memory - macrostructure immediate recall***. Bilateral t-test showed that the initial baseline (T0) difference between the two groups tended to remain at post-intervention (T1) assessment (*t* (29) = 1.99, *p* = 0.056). For ***Text memory - macrostructure delayed recall***, there was a significant main group effect (*F*(1, 2) = 4.80, *p* = 0.039), but no effect of time, or significant group-by-time interaction. There were no significant differences for ***Text memory - microstructure immediate recall*** and ***Text memory - microstructure delayed recall***.

### Generalization measures

No significant between-group differences were found at the baseline assessment on any of the ten dimensions of the ***SEMQ***. There were significant group-by-time interactions on three dimensions: ***Conversations, Slips of attention***, and ***Political & social Events***. The significant group-by-time interaction for ***Conversations*** showed a moderate effect size. Confidence intervals adjusted for *Time since TBI* showed changes in distribution throughout T0, T1 and T2 (95% CIs [-0.1728, 1.2384] [-0.8378, 0.4156] [-0.01142, −1.1234]), indicating some improvement on memorizing conversation content in the experimental group at T1, but showing significance only at the 6-months follow-up (T2). A significant group-by-time interaction was also found for ***Slips of Attention***, with a large effect size. Post-hoc confidence intervals indicated a significant T0-T1 difference (95% CI [−1.07, −0.15]), while the difference between T2 and T0 did not reach significance. Memory for ***Political & social Events*** was significantly increased after the CEP intervention in the experimental group, as indicated by a significant group x time interaction with a large effect size. Confidence interval analysis shows that the effect occurred between T0 and T1 (95% CI [-1.56, - 0.10]), while the difference between T0 and T2 did not reach significance.

There were no significant effects for ***Books & movies, People, Places***, and ***General*** dimensions of the SEMQ. There was a significant main effect of time for ***Use of objects*** (*F*(2, 40) = 12.44, *p* < 0.000) and ***Actions to perform*** (*F*(2, 40) = 4.24, *p* = 0.021), but no significant group effect, nor group × time interaction. No significant effects were found for ***Personal events***, although there was an almost significant group effect (*F*(1, 21) = 4.24, *p* = 0.052). In this case, the control group systematically showed less severe problems on this dimension at the three assessment times.

ANCOVA failed to show statistically significant effects on psychological well-being, as reflected in the **raw scores** of the ***PGWBI***. Additional analyses based on the **clinical thresholds** (well-being, moderate distress, severe distress) for the ***PGWBI*** were performed, and showed clinically meaningful improvements, which are illustrated in Figure 2. Nine (9) participants of the experimental group were clinically classified as severely distressed at baseline (T0) and this number was reduced to 5 after the CEP (T1) and remained relatively stable at 6-months follow-up assessment (T2). Simultaneously, the number of experimental participants classified in the well-being category went from 3 at T0 to 10 at T1 (10/20 participants; 50%) and remained stable six months after (9/17 participants; 53%). In the control group, only one participant progressed toward the well-being category from T0 to T1 and another one at T2. In summary, of the 12 participants having shown improvement, 10 (i.e. 83%) were from the experimental group.

**Figure 2.**
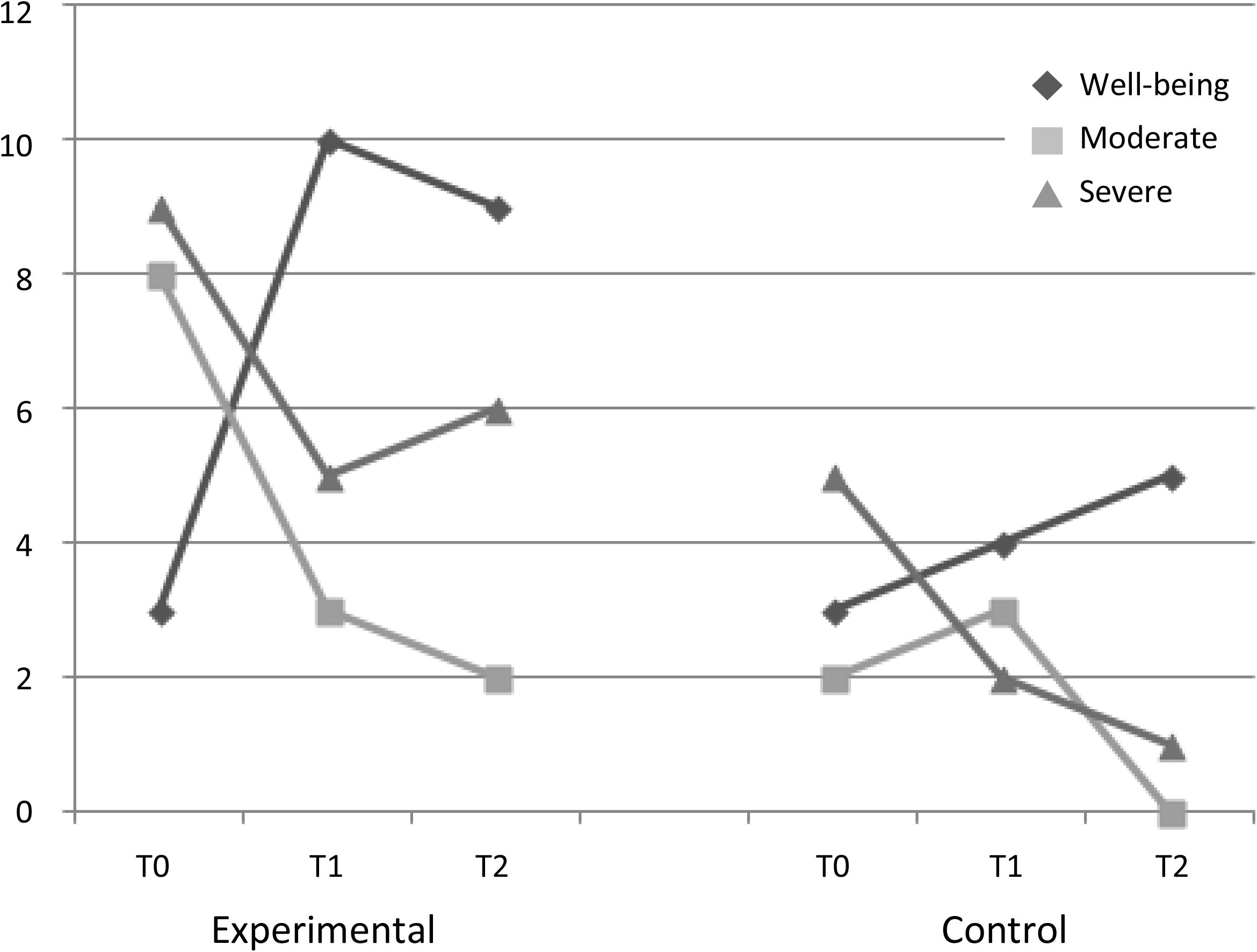
Psychological General Well-Being by clinical category at the three assessment times.

Experimental group participants also completed the ***Satisfaction questionnaire***. The mean score for the question “To which extent did the memory module of the CEP have a positive impact on your daily life?” was 4.00 (*SD* = *0*.*741*) out of a maximum of 5, indicating a perceived positive impact on daily life habits requiring memory abilities. The general satisfaction level for the impact of the entire CEP intervention was 4.75 (*SD* = *0*.*476*).

### Control measures

There were no significant main effects of time, group, nor group-by-time interaction for ***Forward Digit Span***. For ***Backward Digit Span*** there was a significant main effect of time (*F*(2, 40) = 3.33, *p* = 0.04), but no significant main effect of group, nor group-by-time interaction. There was no significant main effect of time, group, nor group-by-time interaction for ***Coding***.

## Discussion

To our knowledge, this is the first study to report on the effects of a multimodal cognitive rehabilitation program developed specifically for older adults with TBI, and to assess its effectiveness for enhancing episodic memory.

### Effectiveness of the CEP on episodic memory

The findings of this study show that the multimodal CEP intervention improves various modalities of episodic memory in older TBI adults that received the training, compared to active TBI controls who received usual rehabilitation focused on resuming daily activities and social roles, but no cognitive rehabilitation. Comparison of baseline measures with post-CEP ones showed a strong group-by-time interaction on one of three primary outcome measures, *Face-name association*, as well as for three dimensions of a real-world generalization measure of memory functioning (SEMQ), *Conversations, Slips of attention, Political & social events*. Experimental participants significantly improved their face-name association abilities and the size of the effect was large. During the CEP intervention, participants learning to associate a person’s name to an interactive image related to a particular feature of the person’s face. In the loci and face-name association methods used in the CEP, the vivid visual imaging representation of a stimulus interacting with other more familiar objects seeks to create novelty at the time of encoding. Such self-initiated novelty has been associated with activation of frontal cortex and its connections, enhancing attention and facilitating encoding, while the presence of multi-modal stimuli boosts simultaneous activation of brain regions associated with verbal (names, words) and visual (facial traits, imagined objects) processing [25]. Our finding of a significant intervention effect on memory of *Face-name associations* and not word lists are compatible with the fact raised by Bottiroli et al. [26] that, contrary to memorizing word lists or reading material, meeting new people is a natural and usual situation where this method, which is directly associated to the context, can be easily practiced.

Three measures of generalization to daily memory functioning were also improved by the CEP: memorizing *Conversations* contents and *Political & Social events*, and *Slips of attention*, showing moderate or large effect sizes. This indicates that these memory dimensions in daily life activities benefited from the learned CEP strategies that aimed at improving the allocation of sustained and selective attention to significant sources of information, such as conversations and news information, with the intention to memorize it; that is, self-enhancing attention during encoding. As we had hypothesized, in addition to improving neuropsychological test performance related to CEP training, the intervention also increased real-world memory function of experimental participants, whereas TBI controls not receiving the CEP did not demonstrate these effects.

The strategies put forward in the CEP target ‘secondary’ memory which is the type of episodic memory often used in daily life situations, where important amounts of information must be retained [27]. Experimental participants thus learned to enhance self-activation when using the memory strategies not only during the training sessions, but also in daily life contexts, where regular practice of the learned strategies constituted a central part of the intervention. They then shared with the group their accomplishments made during the week using the learned methods. Our results thus indicate that older individuals with TBI can benefit from a cognitive rehabilitation program with a strong episodic memory component, such as that put forward by the CEP. Positive impacts of memory training on neuropsychological and self-report memory measures have also been shown in older healthy individuals as well as persons with MCI [8]. It is interesting to note that validation studies using the SEMQ have shown that in healthy aging the *Political and social events* and *Conversation* dimensions items explained most of the variance for the entire measure [28], and that *Conversation* and *Books & movies* were the only items related to actual cognitive deficits in MCI [29]. Increased *Slips of attention* and decreased memory of *Political and social events*, which are the two other SEMQ dimensions that were significantly modified by intervention in our study, could possibly represent an added effect of TBI during aging, as compared to healthy aging or MCI. Those two SEMQ dimensions were previously found to be related to mild TBI in older adults [30], while *Political and social events* and *Books and movies* dimensions were reported as being affected in young adults with mild TBI [31].

With respect to the absence of intervention effects for the other two primary outcome variables, *Word list* learning and *Text memory*, some experimental participants showed difficulties with learning the memory strategies for words and texts. Furthermore, we observed that a few participants had difficulties in learning the reading memory method (FRST). Semantic interpretation, verbal abstract reasoning and verbal synthesis generation problems can be present following TBI. In that respect, Cicerone et al. [32] recommended cognitive interventions for specific language impairments such as reading comprehension after TBI.

Intra-group variability could also have been partly responsible for the absence of significant effects for memory of word lists and texts. Variability of cognitive performance being a characteristic effect of TBI, it could increase the pre-existing inter-individual heterogeneity in cognitive aging [33]. Additionally, contrary to methods of loci and face-name association, the testing condition of memory for texts was dissimilar to the learning one. In fact, participants learned to write a synthesis of main and secondary ideas, create a new title for the story, read their notes and then recalled the story. In the testing situation, participants read a text in silence and then recalled the content orally. It is possible that those differences influenced the results in a TBI sample who may have presented flexibility problems. In this regard, Bottiroli et al. [34] suggested that strategy-adaptation training in cognitive rehabilitation programs for older TBI individuals could help enhance the effects of cognitive rehabilitation. Furthermore, older TBI individuals could probably benefit from training that puts more explicit emphasis on the initiation aspect of using the strategies, since these were not prompted during assessments [32,35].

Since individuals with a more severe TBI have increased difficulties in encoding information, older persons with severe TBI may need a more individualized approach. For instance, a higher number of training sessions could be necessary to develop proficiency in all methods. Some studies reporting effective interventions have invested 15 to 30 hours of training on specific memory strategies [36]. On the other hand, it has been found that encoding words in divided attention conditions is more difficult for older healthy adults [30]. Older persons with severe TBI could then have difficulties in learning and using these strategies because they require dividing attention between cognitive tasks.

### Stability of change over time

The effects of face-name memory were not as stable as hypothesized. Bottiroli et al. [26,34] suggested additional training sessions to facilitate long-term stability of effects of memory training, as well as specific instructions to elders concerning the more suitable circumstances to apply the learned methods. While the CEP intervention explicitly addressed the latter aspect, additional or ‘booster’ sessions and more diversified real-world practice circumstances could have been profitable.

The significant effects shown on the primary outcome and on daily life generalization can be considered as clinically important. However, differences between experimental and control groups changed at the 6-months follow-up assessment. Attrition in the control group at 6-months follow-up could have, in our view, influenced the results. Although there were no significant differences in primary outcomes and memory generalization measures at baseline assessment between the attrition participants and their counterparts who completed the 6-months follow-up assessment in both groups (see Supplementary material), the control group seemed to improve because of the departure of weaker counterparts. Frenette et al. [37] found that mean MoCA scores in the early phases of mild TBI was highly correlated to cognitive functioning, age, schooling, and severity, where individuals with uncomplicated mild TBI obtained higher scores that those with complicated mild TBI. On the other hand, some studies have shown that TBI individuals who succeed in rehabilitation tend to have higher MoCA scores at the beginning than the participants who don’t achieve rehabilitation goals [38]. This seemed to be the case for the control group having completed the study; these participants obtained a mean MoCA score of 26, while attrition control participants had a mean score of 23.67. Otherwise, it is possible that improvement of control group participants was linked to their receiving usual care in the form of rehabilitation interventions focusing directly on daily activities and life habits.

### Generalization to psychological well-being

Psychological factors were not addressed by the CEP interventions. However, we identified potentially clinically important improvements in psychological well-being in experimental participants. Virtually all experimental participants showed an overall improvement in psychological well-being from T0 to T1. While only 3 experimental participants showed a score reflecting well-being status at baseline, this number rose to 10 after the intervention, and this number was stable (i.e., 9) at the 6-months follow-up assessment. Some of the improvements reflected a reduction of psychological distress from severe to moderate. Winocur et al. [39] proposed that self-efficacy could be enhanced by developing strategies that improve cognitive function. Hence, cognitive and memory mastery developed in the experimental group could have had a positive influence on psychological well-being. This was accompanied with a high satisfaction level with regards to positive impacts of memory training, as well as of the overall CEP intervention, on participants’ daily lives. Group experiences also could be an important factor influencing these changes: mutual support and sharing experiences are some of the benefits of group participation, in addition to the already demonstrated cognitive benefits. Considering that depression is frequently seen following TBI even if age still a controversial risk factor [40], group interventions may represent a promising approach to influence mood through a cognitive training.

### Study limitations

The present study has some limitations that should be taken into consideration when interpreting our findings. It is impossible to determine if only the memory module of the CEP intervention is related to the observed changes in the experimental group. Such a multimodal approach to cognitive rehabilitation is warranted clinically and is evidence-based considering the important variability in the aging process and clinical manifestations of TBI. However, the fact that intervention effects were found on tests measuring memory abilities that were targeted by the memory module of the CEP suggests that there was a direct effect of memory intervention. Also, the small size or our sample, with the largest proportion of participants having a mild TBI, does limit the scope of conclusions, and we have also pointed out earlier the possible influence of attrition on the results. However, effects sizes of significant changes were moderate to large, indicating that these were strong effects that were related to the memory interventions. Our experimental group received the CEP intervention as well as usual care (holistic rehabilitation interventions without cognitive rehabilitation) while active controls also received usual care, albeit without any cognitive rehabilitation. There thus remains the possibility that some effects of CEP interventions were masked by the positive influence of this holistic rehabilitation approach. At the same time, we considered that any significant change favoring the experimental group would mean that interventions effects were more robust.

## Conclusions

Traumatic brain injury in older adults is a public health problem needing urgent attention from scientific and clinical communities as well as from stakeholders. Yet, rehabilitation programs developed for older TBI individuals are sparse or inexistent. We have tailored a new cognitive rehabilitation program, the CEP, integrating cognitive strategies that have been shown effective in other populations (normal aging, younger TBI, MCI) to target common cognitive problems associated with those conditions. Our results show that CEP intervention can improve neuropsychological and self-reported measures of episodic memory (namely face-name associations and self-reported improvement of memory for conversations and political and social events, as wells as reductions in slips of attention). Clinical improvements of psychological well-being were also observed as a possible generalization effect, since self-efficacy is enhanced by cognitive improvement in daily life activities. Considering these clinically important results, the accessibility to rehabilitation for older people who have sustained a TBI must be closely monitored and increased.

## Supporting information

ICMJE Form

## Data Availability

Data could be made available upon request.

## Acknowledgments

We wish to thank the TBI Programs of the Lucie-Bruneau and Le Bouclier Rehabilitation Centres, as well as the Trauma Program of the MUHC for their important role in participant recruitment. We also thank the individuals who participated in this research, sharing generously their time, efforts and energy. We would like to pay our gratitude and our respects to Dr. Donald T. Stuss, who, sadly, passed away in September 2019, and to Drs Gordon Winocur and Brian Levine for advice and encouragement at the early stages of this study. We thank Webneuro from Université de Lausanne in Switzerland for authorizing the use of some of the neuroanatomical representations featured in CEP manuals. We are indebted to Véronique Beauséjour for the enormous work of assessment, as well as Guylaine Bélizaire, Michel Ouellette, Christel Cornelis and Alexandra F. Girard who also acted as research assistants for this project. We acknowledge the statistical work and advice of Miguel Chagnon. This project was funded by the Fonds de recherche du Québec – Santé (FRQS; research grant 22319 to M.M. and scholarship to E.C.), the Quebec Rehabilitation Research Network (research grant 09-10DS-06 to M.M.), the Lucie-Bruneau Rehabilitation Centre (research grant to E.C. and M.M., and clinical research hours to E.C.). The authors have no conflict of interest.

## Supplementary material

## Material and methods

### Additional information on participants

Inclusion criteria were: a) diagnosed with mild, moderate or severe TBI at least six months before enrolment in study, based on the World Health Organization criteria [16]: *Mild*: length of loss or altered level of consciousness (LOC) 0-30 minutes, Glasgow Coma Scale (GCS) score 13-15/15, negative or positive brain imaging (CT-Scan or MRI), post-traumatic amnesia (PTA) duration <24 hrs; *Moderate*: LOC 30 min-24 hrs, GCS score 9-12, positive brain imaging, PTA duration 1-14 days; *Severe*: LOC >24 h, GCS score 3-8, positive brain imaging, PTA duration >2 weeks [16]; b) post-traumatic amnesia period must be already resolved; c) aged at least 55 years; d) fluent in French (speaking, understanding, reading). Exclusion criteria were: a) previously received or receiving another specific or direct cognitive intervention focusing on similar or identical cognitive functions; b) diagnosis or documented clinical impressions of dementia (medical files) or MOCA score lower than 20; c) diagnosis of an active psychiatric condition; d) consumption of alcohol (drinking 5 or more drinks on the same occasion on each of 5 or more days weekly in the past 30 days), or consuming illicit drugs.

This study was conducted (from recruitment to the 6-months follow-up) between September 2012 and April 2015. Out of the 42 referred individuals assessed for eligibility, 37 gave consent to participate either as an experimental or control participant, as would be determined by the allocation procedure. By order of reference, participants were then assigned semi-randomly to experimental or active control sub-groups of five, and TBI severity was matched across sub-groups. At the end of the recruitment period, 23 participants had been assigned to the experimental group, and 14 had been assigned to the control group. Three participants of the experimental group and two control subjects withdrew from the study. Baseline (pre-intervention - T0) evaluations were performed with 20 participants from the experimental group, who received the CEP intervention plus usual care, as well as with 12 active control participants, who received usual care and were encouraged to keep cognitively active in addition to participating in study assessments. Usual care was defined as interventions within a holistic interdisciplinary rehabilitation program focused on resuming daily activities and social roles, if needed, as determined by treating physician, and did not include any form of cognitive rehabilitation. Three (3) participants in the experimental group missed one intervention session each, and they were individually delivered by the experimenter. There were missing data on some measures for a small number of participants in both groups (e.g., non-completion of questionnaires by participants, fire alarm during testing).

### Content and structure of the Cognitive Enrichment Program (CEP)

| ***First module – Introduction and Self-awareness*** |  |
| --- | --- |
| Session 1 | Introduction to the CEP. |
| Session 2 | Impact of aging on brain and cognitive functions. Brain plasticity during aging. |
| Session 3 | TBI and aging. Cognitive intervention during normal aging and rehabilitation for ageing persons with neurological conditions. |
| Session 4 | Self-awareness, self-assessment of cognitive strengths and difficulties. |
| ***Second module – Attention and Memory*** |  |
| Session 5 | Attention and specific attention modalities used in daily life. Selective and divided attention exercises. |
| Session 6 | Familiarisation with different types of memory, and main phases of episodic memory in relation with encoding. |
| Session 7 | Introduction to MEMO. Global assessment of participants' ability to create interactive visual images. Practicing generation of interactive images. |
| Sessions 8, 9 & 10 | Three MEMO memory approaches: the method of loci and face-name associations, both using visual imagery mnemonics, and the FRST method to memorize short texts using semantic associations. |
| Session 11 | Review of attention and memory strategies. |

| ***Third Module – Executive Functions*** |  |
| --- | --- |
| Session 12 | Definition, examples, and executive functions self-assessment questionnaire. |
| Session 13 | Daily attention difficulties and executive dysfunctions. |
| Session 14 | General planning method. |
| Sessions 15 & 16 | Planning and organizing a personal plan. |
| Sessions 17, 18 & 19 | Strategies for solving problems. Identification of a problem. Dividing a problem into smaller problems. Generating plans, pondering alternatives and choosing solutions, identification of steps, resources and time. Solving fictive and personal daily life problems. |
| Session 20 | Introduction to Goal Management and self-regulation methods. Inattention. Absentmindedness versus mindfulness as crucial dimensions against or for self-regulation. The Goal Diagram. |
| Session 21 | Awareness exercises for inattention and automatisms. Introduction to the STOP technique. Defining explicit goals and strategies. Application of problem-solving strategies to identify steps and partial strategies. |
| Session 22 | Practice of Stop-and-state technique. Strategies for keeping in mind main goal and main strategy. Application of visual imagery. |
| Session 23 | Self-assessment using the Stop-and-state technique for reviewing strategies and results. Fictive and personal daily life exercises of goal management and self-regulation. |
| Session 24 | Global review of the three modules. CEP appraisal by participants. |

### Intervention delivery

At each session, participants were presented with the conceptual framework for the specific cognitive strategies to be targeted, performed practical exercises, and were assigned homework. Homework was reviewed by the experimenter to give feedback, identify and resolve errors, and give supplementary explanations. The main teaching and intervention techniques were practical theoretical explanations of the general basis of each strategy in association with TBI, typical memory problems, descriptions of the steps of each method, modeling the use of techniques, facilitating practice of techniques in a progressive manner (e.g., gradually increasing number of words to memorize, choosing meaningful texts to read and memorize), detailing daily situations in which the techniques are more suitable to be used (situations serving also as a context for practice homework), and group discussions for revising homework. An intervention manual for the CEP was developed to insure reliability and reproducibility of interventions. A participant’s manual was built by including material and notes relating to the various approaches, strategies, exercises, and homework. A section of the notebook was reserved to writing personal notes and comments by the participant. This notebook included a calendar and a model of a personal organizer.

### Additional information on assessments and measures

There were three time-points for assessments: pre-intervention baseline (T0), that is one week before starting the CEP for experimental participants; 14-weeks (post-intervention – T1), that is approximately one week after end of CEP intervention for the experimental group and 14 weeks following baseline assessment of the control group; 6-months post-intervention (follow-up – T2), that is 6 months after the T1 assessment for both groups. When available, different equivalent versions of the same tests were used in counterbalanced administrations in order to diminish practice effects. To reduce fatigue or interference effects, the order of administration by type of cognitive function assessed (attention, memory, executive functions) was also counterbalanced between and within assessment sessions. With the aim of reducing fatigue during testing, self-report questionnaires were filled by participants at home between sessions.

### Primary outcome measures

For face-name association, participants had to associate 12 previously learned face-name pairs, and for word lists, 12 previously learned words had to be recalled. For word lists and text memory, measures of immediate and delayed recall were obtained. For memory of texts, results were divided into 23 macrostructure elements (main ideas of the text giving a meaning to the story), and 24 microstructure elements (specific details about the story). A slight difference with Belleville et al.’s [12] administration procedure is that at the follow-up assessments we did not instruct our participants to use the mnemonic strategies learned during the program. We wished to measure spontaneous utilization of strategies learned, in order to see if participants would be able to self-initiate the use of learned mnemonic strategies.

### Generalization measures

SEMQ questions are answered using a Likert scale (1 to 6; never to always), where better perceived memory performance is noted by a lesser score. The PGWBI is a self-report questionnaire measuring self-perceived well-being. This measure is obtained through items assessing psychological distress, and by items reflecting psychological well-being. This dual approach allows this instrument to obtain a very robust measure of the construct. The scale consists of 22 self-administered items, rated on a 6-point scale, which are summarized into a global maximum summary of 110 points, representing highest attainable level of well-being. The Satisfaction questionnaire used a Likert scale (0-5, no impact to high impact).

### Control measures

Forward and Backward Digit spans and Coding subtests from the WAIS-III were used since the CEP does not target cognitive functions measured by those tests: attention span, working memory, visual selective attention, motor and cognitive processing speed. A score lower than 20 on the MoCA initiated a decisional process to eliminate or not a participant by performing a detailed medical file analysis, and by exploring if there were any errors on the Orientation task of the MoCA. WAIS-III Vocabulary, reputed to measure crystallized intelligence, was used as an indicator of premorbid level of intelligence.

## Results

### Primary outcome measures statistics for non-significant results

*Face-name association*: pre-post-intervention difference not maintained at the 6-months follow-up (95% CI [-0.77, 4.60]).

*Word-list - Immediate recall*: no significant main effect of group (*F*(1, 22) = 0.81, *p* = 0.377).

*Word-list - Delayed recall*: no significant effect of group (*F*(1, 21) = 2.23, *p* = 0.151).

*Text memory - Macrostructure immediate recall*: no significant main effect of time (*F*(2, 44) = 0.13, *p* = 0.877), or group (*F*(1, 2) = 2.56, *p* = 0.124).

*Text memory - Macrostructure delayed recall*: no significant main effect of time (*F*(2, 44) = 0.10, *p* = 0.908).

*Text memory - Microstructure immediate recall*: no significant main effect of time (*F*(2, 44) = 0.64, *p* = 0.5324), or group (*F*(1, 22) = 0.27, *p* = 0.608).

*Text memory - Microstructure delayed recall*: no significant effect for time (*F*(2, 44) = 0.27, *p* = 0.765), or group (*F*(1, 22) = 0.27, *p* = 0.609).

### Generalization measures statistics for non-significant results

*Books & movies, People, Places*, and *General* dimensions of the SEMQ: no significant main effect of time, or group (respectively, Books & movies: *F*(2, 40) = 0.82, *p* = 0.45; *F*(1, 21) = 0.02, *p* = 0.90; People: *F*(2, 40) = 0.01, *p* = 0.99; *F*(1, 21) = 0.06, *p* = 0.80; Places: *F*(2, 40) = 0.01, *p* = 0.991; *F*(1, 21) = 3.41, *p* = 0.079; General: *F*(2, 40) = 0.15, *p* = 0.857; *F*(1, 21) = 2.57, *p* = 0.124).

*Use of objects*: no significant effect of group (*F*(1, 21) = 0.28, *p* = 0.599).

*Actions to perform*: no significant effect of group (*F*(1, 21) = 1.41, *p* = 0.2461).

*Personal events*: no significant effect of time (*F*(2, 40) = 0.51, *p* = 0.601).

*PGWBI*: no significant effect of time (*F*(2, 36) = 1.39; *p* = 0.262), or group (*F*(1, 18) = 1.66; *p* = 0.213).

### Control measures statistics for non-significant results

*Forward Digit Span*: no significant main effect of time (*F*(2, 40) = 0.02, *p* = 0.99) or group (*F*(1, 21) = 0.07, *p* = 0.79).

*Backward Digit Span*: no significant main effect of group (*F*(1, 20) = 0.30, *p* = 0.59).

*Coding*: no significant main effect of time (*F*(2, 44) = 0.03, p = 0.97), or group (*F*(1, 22) = 0.48, *p* = 0.49).

### Statistics for completed/attrition participants

Control participants lost before the 6-months follow-up assessment were compared to control counterparts who completed the study. Comparing the completed/attrition controls (n= 7/5) at baseline assessment, we observed that although attrition controls were slightly older, their TBI was more recent, they presented more vascular risk factors, and had lower psychological general well-being and lower scores on some of the primary outcome measures (i.e., 2 of 7 memory scores, and 7 of the 10 dimensions of the SEMQ), these differences did not reach significance except for one memory variable, *Text memory - macrostructure immediate recall* (15.67 > 10.67; *t* (10) = 2.43; *p* = 0.035). On the other hand, experimental attrition participants showed fewer problems than their counterparts on 2 of 7 memory scores, although not reaching significance. Furthermore, no significant differences were found between experimental completed/attrition in terms of age, vascular burden, time since TBI or *PGWBI*.

## TREND Statement Checklist

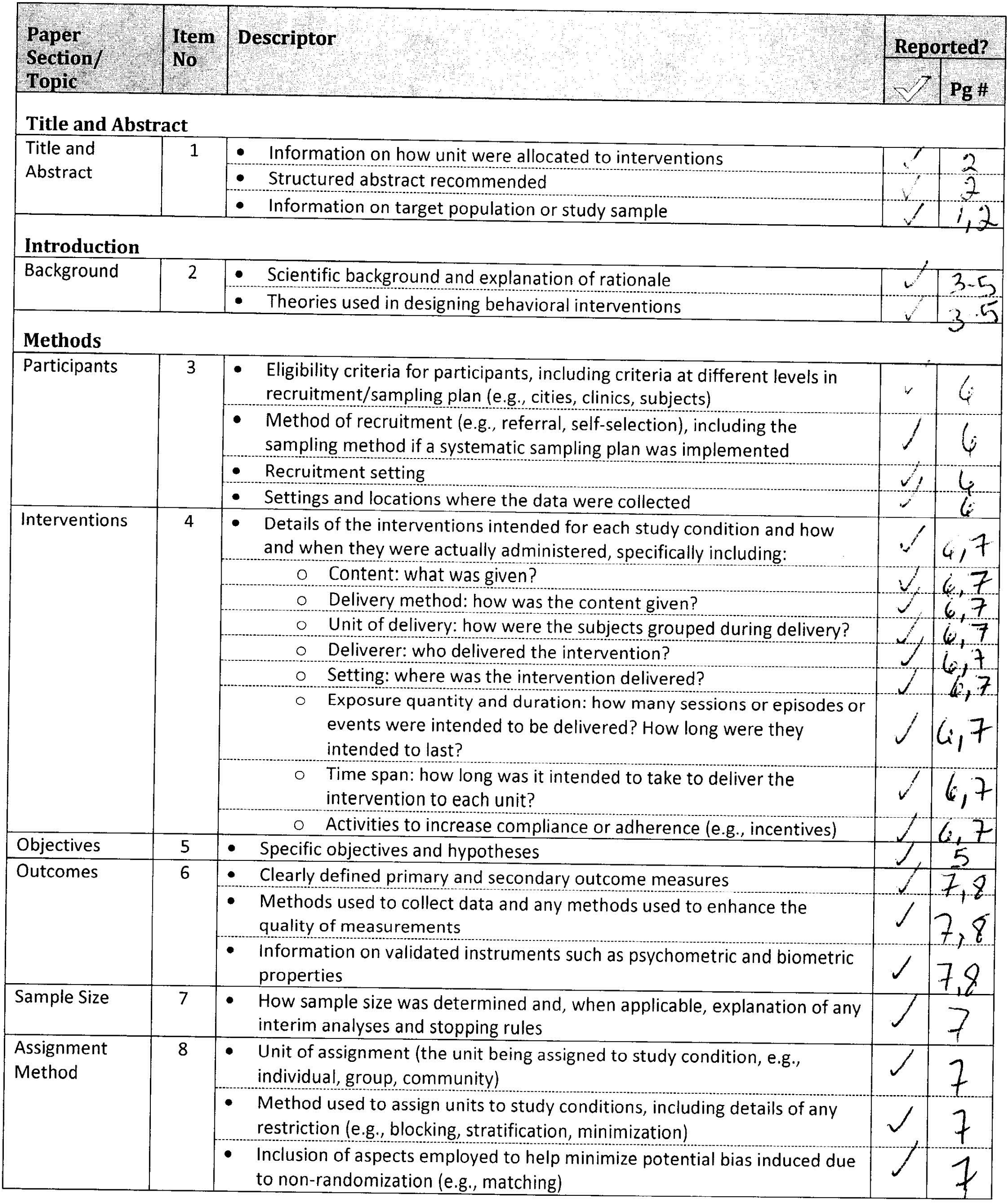
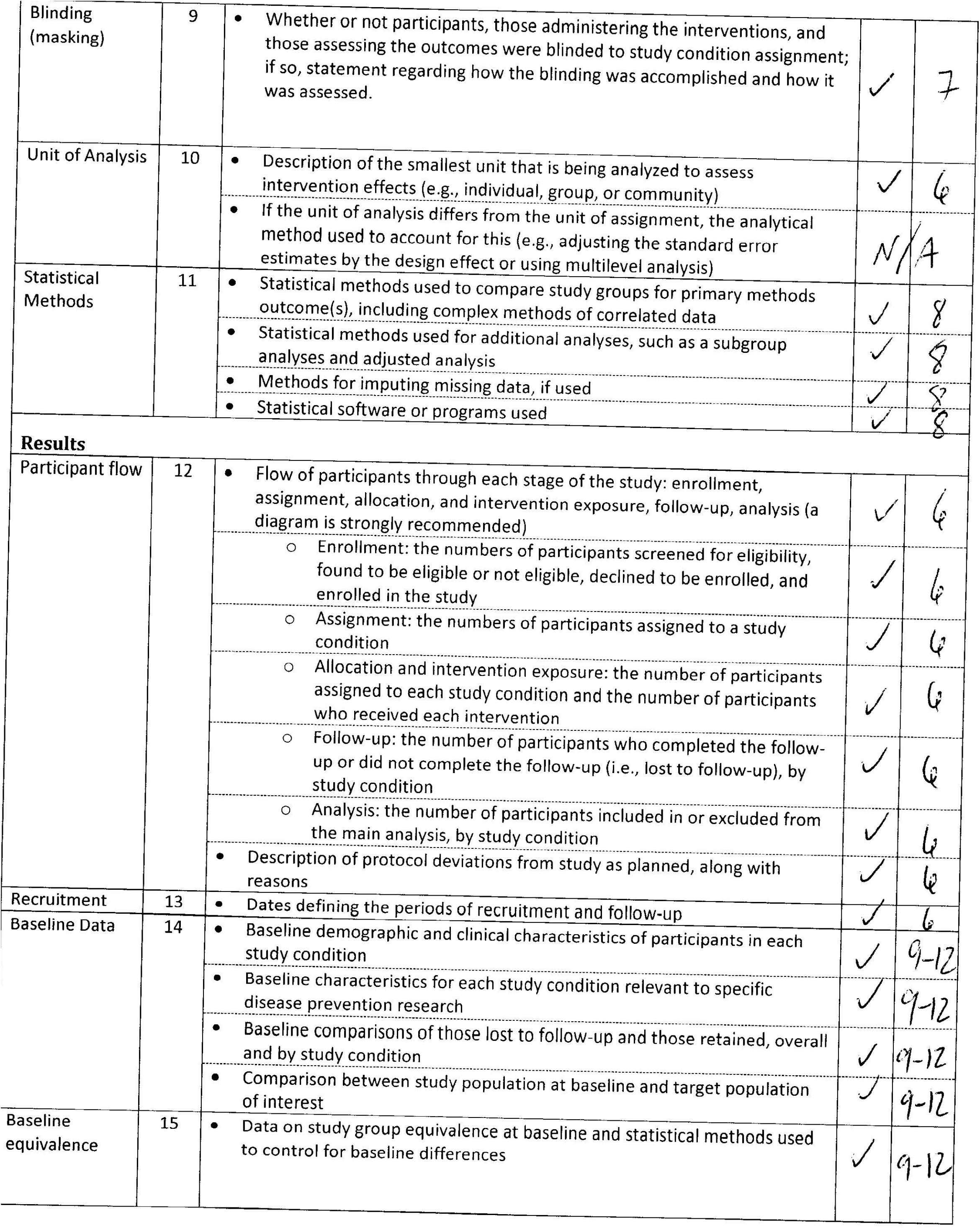
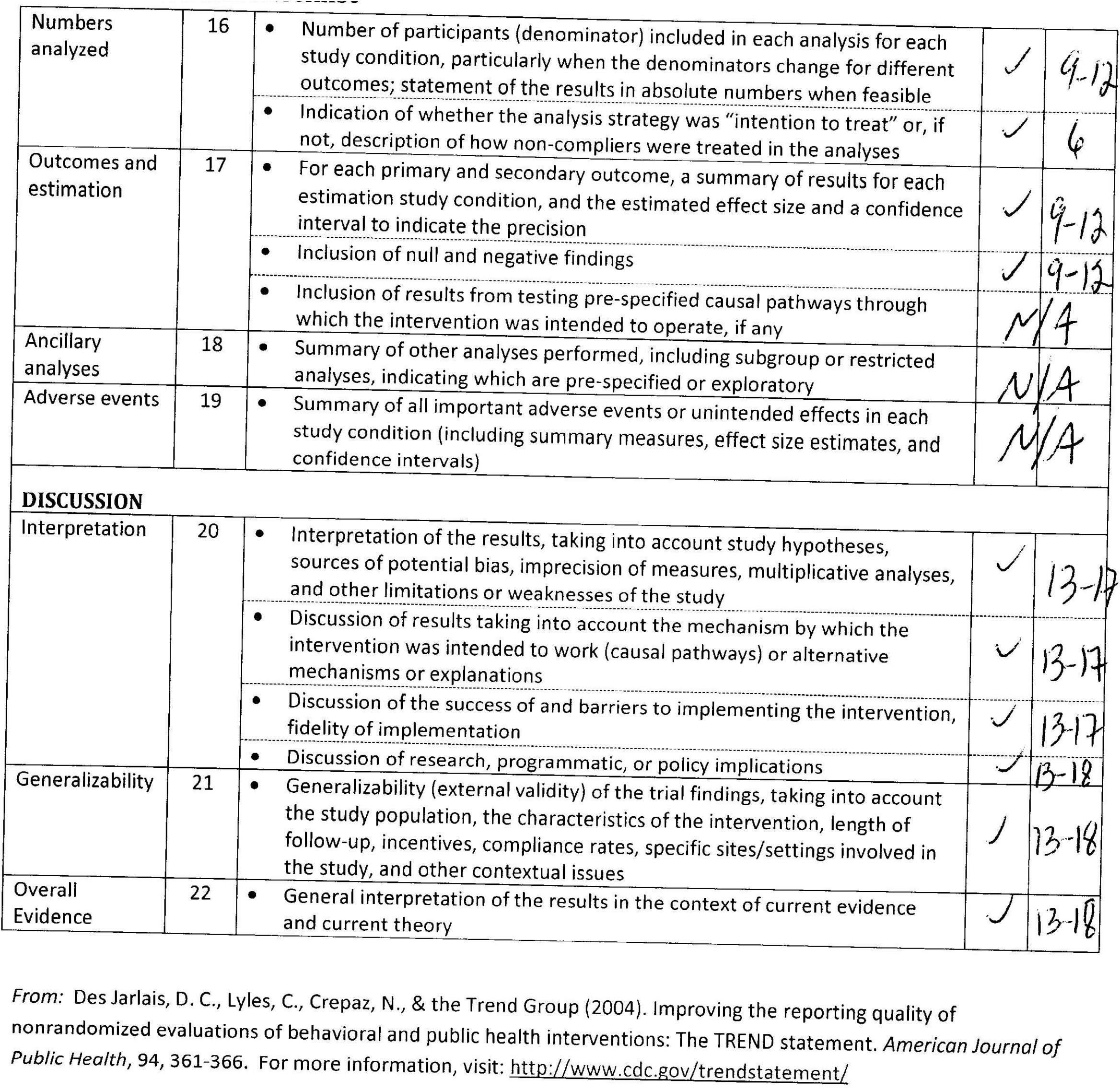

